# Machine learning and burden analyses highlight novel candidate genes in Parkinson’s disease

**DOI:** 10.64898/2026.01.22.26344646

**Authors:** Sitki Cem Parlar, Eric Yu, Sajanth Kanagasingam, Zhao Zhang, Lang Liu, Morvarid Ghamgosar Shahkhali, Cloe Chantereault, Nathan Karpilovsky, Donovan Worrall, Rhalena A Thomas, Konstantin Senkevich, Edward A Fon, Ziv Gan-Or

**Affiliations:** Department of Human Genetics, McGill University, Montréal, Québec, Canada; Montreal Neurological Institute-Hospital (The Neuro), McGill University, Montréal, Québec, Canada; Department of Neurology and Neurosurgery, McGill University, Montréal, Québec, Canada; DataTecnica LLC, Washington, DC, USA; Early Drug Discovery Unit (EDDU), Montreal Neurological Institute-Hospital (The Neuro), Montreal, QC, Canada; Department of Specialized Medicine, Division of Medical Genetics, McGill University Health Centre, Montreal, Québec, Canada

**Keywords:** Parkinson’s disease, Machine learning, Rare variants, Burden analysis, Gene prioritization

## Abstract

Genome-wide association studies (GWAS) have identified numerous risk loci for Parkinson’s disease, yet identifying specific causal genes remains a major challenge due to non-coding associations and complex linkage disequilibrium. Here, we present a systematic framework integrating machine learning-based gene prioritization with high-resolution rare variant burden analysis. Using an XGBoost machine-learning model trained on 285 multi-omic features, including brain-specific eQTLs and single-cell expression, we prioritized 406 out of 3,105 genes across 147 risk loci. The list of prioritized genes underwent gene- and domain-level rare variant burden analysis via optimal sequenced kernel association test (SKAT-O) across the Accelerating Medicines Partnership for Parkinson’s Disease and UK Biobank cohorts (N case = 6,435, N proxy = 13,889, N control = 343,160). Meta-analysis of rare variants at the gene and domain levels replicated established associations (*GBA1*, *LRRK2*) and identified six novel potential risk genes *ANKRD27* driven by p.Arg21Cys, *UBXN2A*, *FAM171A1*, *ERCC8*, *BNC2*, and *ADNP*. Domain-level analysis uniquely uncovered a significant association in the zinc-finger domain of *ADNP*, which was masked in gene-level tests. Additionally, at cohort-level, one novel gene was nominated: *LRRC45* driven by p.Met607ArgfsTer57. Collectively, our results indicate that integrating machine-learning prioritization with gene- and domain-level burden testing identifies novel genes potentially involved in Parkinson’s disease, with further validation needed to elucidate causality and mechanisms.

## Introduction

Parkinson’s disease (PD) is the fastest-growing neurodegenerative disorder globally, affecting millions of individuals and imposing a substantial burden on healthcare systems.^1^ While approximately 15% of cases have a family history of PD, the genetic architecture of the disease remains largely elusive, with a significant portion of heritability unexplained. ^2^ Large-scale genome-wide association studies (GWAS) have been instrumental in mapping the genetic landscape of PD, revealing numerous risk loci. However, the vast majority of these signals map to non-coding regions, making the identification of the specific causal variants, genes and mechanisms within these loci a major challenge.

To bridge the gap between statistical association and biological function, novel computational strategies are required. Recently, we demonstrated that machine learning approaches integrating multi-omic data can effectively prioritize high-confidence candidate genes within GWAS loci.^3^ By leveraging tissue-specific transcriptomic and epigenetic features, these models move beyond simple genomic distance to identify drivers of disease risk. Furthermore, GWAS signals primarily capture common variants with small effect sizes, potentially neglecting the contribution of rare, highly penetrant variants.

Due to limited statistical power, rare variants are conventionally analyzed via burden testing, which aggregates variants into a single “gene-level” unit. While effective for well-established genes such as *GBA1*, *LRRK2*, and *SNCA*, this approach lacks functional resolution. Pathogenic mechanisms are often specific to distinct structural regions of a protein, such as the kinase domain of *LRRK2*,^4^ meaning that aggregating variants across the entire gene may dilute potent signals with benign noise. Consequently, the current list of validated PD targets remains small, limiting our ability to uncover novel therapeutic targets.

In this study, we combined machine-learning-based gene prioritization on the most recent European PD GWAS study, which is the largest of its kind, with a high-resolution rare variant burden analysis. By demarcating proteins into their constituent functional domains, we aimed to increase the specificity of genetic association testing. This dual approach allows us to not only refine the list of prioritized candidates from European GWAS loci but also to uncover novel, domain-specific pathogenic mechanisms that are missed by traditional gene-level analyses.

## Methods

### Machine Learning and Prioritization Framework

#### Study Overview

Figure 1 illustrates the machine learning framework and analytical workflow used in this study.

**Figure 1.**
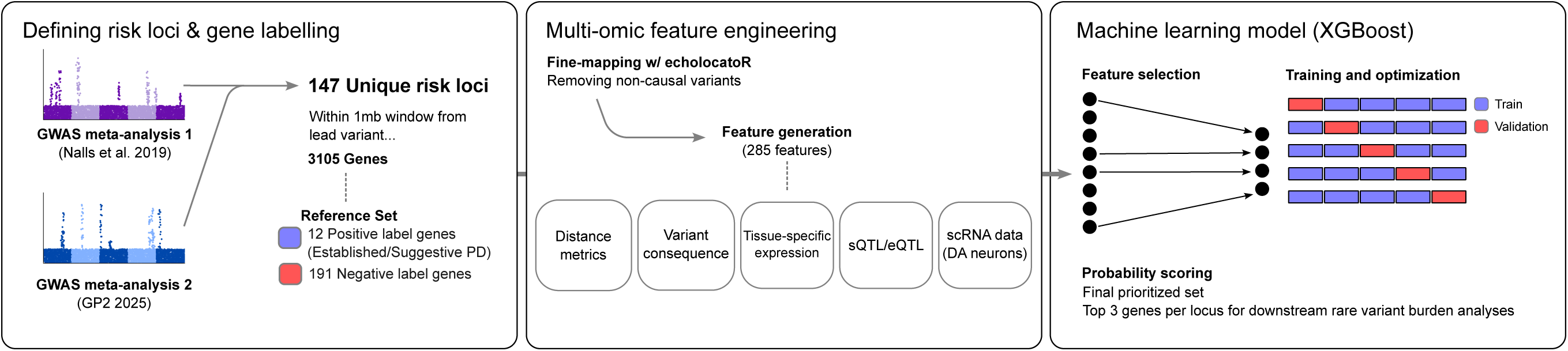
Workflow for machine learning-based prioritization of PD risk genes. Risk loci were defined from GWAS meta-analyses to create a labeled training set **(Left)**. Genes were annotated with fine-mapped multi-omic features **(Middle)** and used to train an XGBoost model with 5-fold cross-validation to identify high-probability candidates **(Right)**.

#### Definition of Loci and Genes

To capture a comprehensive and high-yield map of the genetic architecture underlying Parkinson’s Disease (PD), we constructed a composite set of risk loci by integrating the two largest available genome-wide association study (GWAS) meta-analyses.^5,6^ We incorporated summary statistics from the recent European GP2 study, which identified 134 risk loci, and manually reinstated 13 unique loci from the previous meta-analysis by Nalls et al. (2019) that were absent in the newer release. This integration resulted in a final dataset of 147 unique PD risk loci. Following the definitions in the cited GWAS studies, significant variants within a 250kb window were merged to define distinct risk loci. For our analysis, all protein-coding genes (N = 3105) located within a +/- 1mb window of the independent GWAS lead variants within each locus were extracted.

For our machine-learning based prioritization model, we relied on a set of genes labeled as positive and negative. This reference set included the original seven well-established PD genes (*GBA1, LRRK2, SNCA, GCH1, MAPT, TMEM175, VPS13C*)^2^ alongside five additional genes whose associations provide limited evidence (*SCARB2, CTSB, GRN, GALC, GPNMB*).^7–10^ The remaining 191 genes within the risk loci from which the 12 positive labelled genes were taken were labeled as negative.

#### Multi-omic Features

To filter for potential causal variants, we utilized echolocatoR,^11^ a Bayesian fine-mapping pipeline, incorporating the resulting credible sets and independent risk signals directly into the feature generation process. We characterized each gene using a high-dimensional set of 285 features derived from genomic, transcriptomic, and epigenomic data mapped to the hg19 reference genome. The list of 285 features and the sources from which they were accessed are detailed in the Supplementary Tables 1 and 2 respectively. Briefly, recognizing that physical proximity is often a strong predictor of causality,^12^ we derived composite distance metrics (including minimum and mean distances), calculated from the set of risk variants, comprising independent GWAS lead variants and high-confidence causal candidate variants identified by echolocatoR, to both the gene body and the transcription start site. Variant severity was quantified using scores from Ensembl Variant Effect Predictor (VEP)^13^ and PolyPhen-2.^14^ To capture relevant biological context, we utilized the Functional Mapping and Annotation (FUMA) platform^15^ to map variants to genes via expression and splicing quantitative trait loci (eQTLs/sQTLs) using the UK Biobank release 2b 10k European reference panel. Furthermore, we integrated 3D chromatin interaction data and tissue-specific expression profiles from FANTOM^16^ and GTEx,^17^ while cell-type resolution was achieved using single-nucleus RNA sequencing (snRNA-seq) data from dopaminergic neuron subpopulations.^18^

To standardize features across heterogeneous loci, we applied a “neighbourhood scoring” method.^12^ Continuous features were scaled relative to the maximum value observed within that specific locus, ensuring the model prioritized local signal strength rather than absolute global values. Distance metrics underwent negative log transformation to invert their relationship with the score, ensuring that closer proximity resulted in a higher prioritization value.

#### Machine Learning Model

We implemented an XGBoost gradient boosting model^19^ to distinguish genes with high-probability of being associated with PD within the loci. The model was trained on the defined positive and negative sets. To correct for class imbalance, we set the scale_pos_weight parameter to the ratio of negative to positive labels. Training proceeded in a two-step phase; an initial feature selection pass removed redundant variables from the starting pool of 285 features to isolate the most predictive subset, followed by the training of the final model on this refined set of 48 features. We optimized hyperparameters using 5-fold cross-validation and maximized the score of correct positive predictions by using mean average prediction as an evaluation function. For every gene in the 147 loci, the model generated a probability score (Supplementary Table 3).

The top three scoring genes per locus were then prioritized for our downstream analyses to be assessed for their association to PD.

### Gene-Level and Domain-Level Rare Variant Burden

#### Study Population

Our study population was drawn from two large whole-genome sequencing (WGS) cohorts: Accelerating Medicines Partnership for Parkinson’s Disease (AMP-PD)^20^ and UK Biobank (UKBB).^21^ To assess rare variant burden, we performed two distinct meta-analyses to balance phenotypic rigor with sample size (Table 1).

**Table 1.**
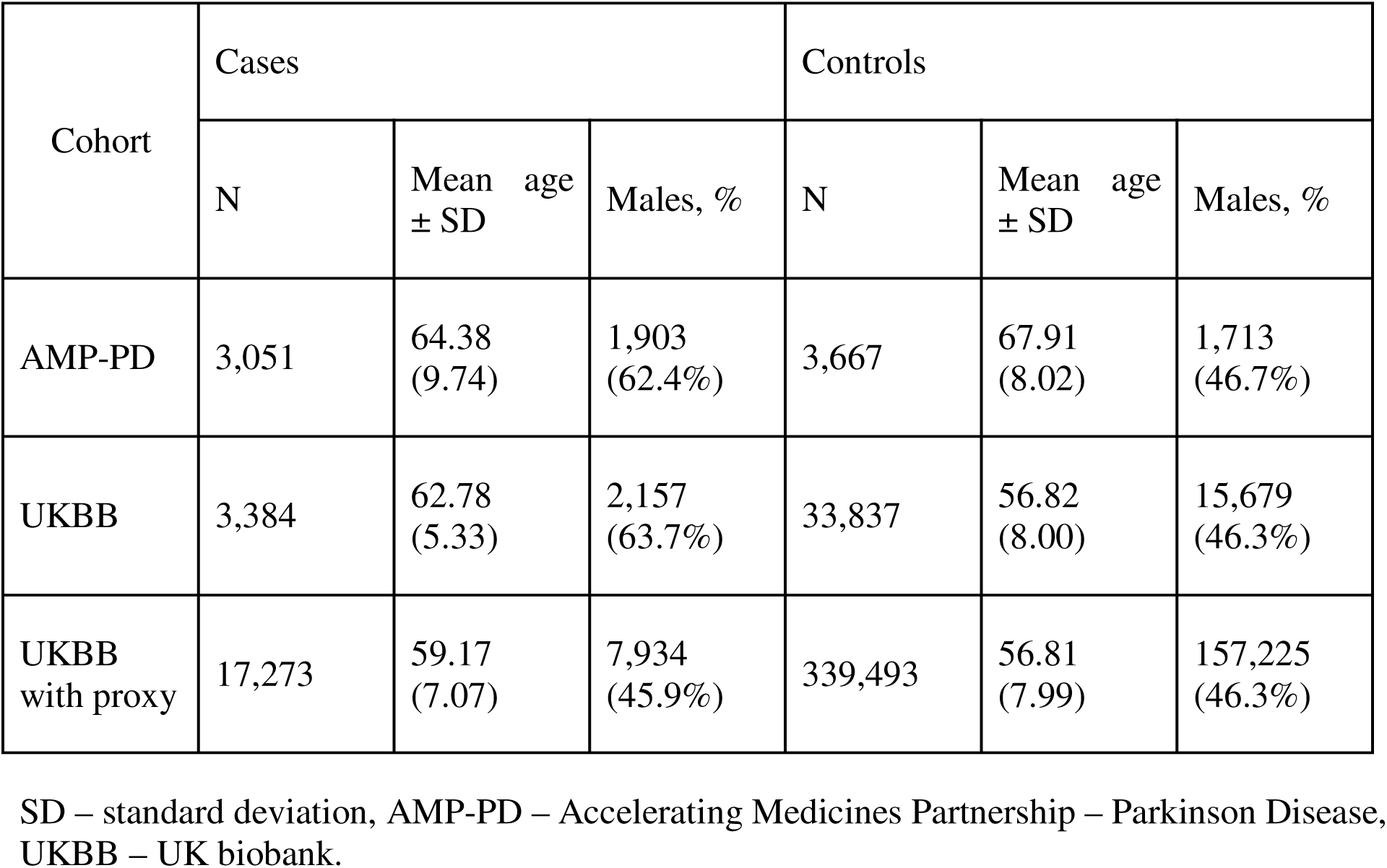
Study population demographics for rare variant burden analysis.

First, to prioritize clinical specificity, we meta-analyzed the AMP-PD cohort (N cases = 3,051, N controls = 3,667) with a subset of the UKBB containing PD cases identified via clinical diagnosis, self-report, or death registry records, matched with 10x randomly sampled controls (N cases = 3,384, N controls = 33,837). This primary clinical meta-analysis encompassed a total of 6,435 cases and 37,504 controls.

Second, to maximize statistical power, we performed a broad meta-analysis combining AMP-PD with the entire UKBB dataset that included both PD cases and proxy cases (N cases = 17,273, N controls = 339,493). The proxy cases were defined as participants self-reporting a parental or sibling history of PD (UKBB Data-Fields 20107, 20110, and 20111). This yielded a total cumulative sample size of 20,324 cases and 343,160 controls.

Regarding diagnostic standards, AMP-PD utilized the established Movement Disorder Society (MDS) criteria,^22^ while UKBB clinical diagnoses were based on the UK Parkinson’s Disease Society Brain Bank (UKPDSBB) criteria.^23^

#### Sequencing and Quality Control

Whole-genome sequencing was performed using the Illumina NovaSeq 6000 (UKBB)^21^ and HiSeq X Ten (AMP-PD)^20^ platforms, aligned to the hg38 reference genome. We performed quality control steps at both sample and genetic level. For the sample-level quality control, we filtered samples to ensure population homogeneity and unrelatedness. For the UKBB cohort, exclusions were made based on provided quality control fields. We retained only individuals of European ancestry as defined by genetic ethnic grouping (Data-Field 22006) and removed related individuals to ensure sample independence. Additionally, we excluded samples flagged for sex chromosome aneuploidy or identified as outliers for missingness. The AMP-PD cohort was already quality controlled as previously described.^20^ Consequently, we restricted the sample list to unrelated individuals classified as “White” based on cohort demographics.

Following sample selection, we applied strict genetic-level filtering using PLINK 2.0^24^ and bcftools 1.16.^25^ For the UKBB cohort, WGS data was obtained through the UK Biobank Research Analysis Platform (DNA-nexus). In addition to the upstream quality control performed by the source institution, variants were retained if they met Genotype Quality (GQ) > 25, Read Depth (DP) > 25, and missingness rate < 5%. For the AMP-PD cohort, as GQ data were not available, filtering relied on site-level DP and missingness. Variants were retained if they had a DP > 25 and a missingness rate < 5%. Further, in both cohorts, we filtered for rare variants by applying a minor allele frequency threshold of ≤ 0.01.

#### Gene and Domain Demarcation

For the list of genes used for the machine learning prioritization, the canonical GENCODEV48 coordinates were obtained via UCSC’s Table Browser.^26^ To resolve transcript discrepancies, we prioritized MANE Select transcripts, as they represent a clinically standardized and biologically relevant reference.^27^

To define protein domains, we developed an automated pipeline integrating the Ensembl REST API^28^ with InterPro^29^, targeting the previously identified MANE Select transcripts. We queried the API to extract protein features, relying exclusively on three databases selected for their extensive manual curation and complementary biological focus: Pfam-A, SMART, and PROSITE Profiles.^30–32^ Furthermore, we strictly filtered for features possessing a validated InterPro Accession (IPR ID). This requirement ensures that each feature represents a consensus signature integrated across the InterPro member databases, providing a high-confidence set of annotations.^29^

To ensure discrete domain representation and avoid redundancy, we applied a two-step refinement process. First, we addressed fragmented alignments using a “Consensus Range” logic, where discontinuous fragments or tandem repeats sharing a unique InterPro ID were collapsed into a single inclusive span (absolute start to absolute end). Second, to resolve overlaps between distinct domain types, we applied a strict hierarchy (Pfam > SMART > PROSITE). Lower-priority domains overlapping a higher-priority entry by more than 20 amino acids were discarded, ensuring that no variant was double-counted across multiple domain tests. Finally, the validated amino acid boundaries were mapped to hg38 genomic coordinates.

#### Variant annotation

Functional annotation of variants was performed using the Ensembl Variant Effect Predictor (VEP) software v114.0,^13^ referencing the GENCODE V48 gene set.^26^ Variants were assigned Phred-scaled scores based on predicted deleteriousness using the Combined Annotation Dependent Depletion (CADD) v1.7 framework.^33^ Additionally, population-specific allele frequencies were extracted from The Genome Aggregation Database (gnomAD).^34^

Following annotation, variant-level allele frequencies and counts were produced using PLINK v1.9.^35^ To evaluate individual variant associations, Odds Ratios (OR) and 95% Confidence Intervals (CI) were calculated using 2x2 contingency tables. We reported both unadjusted ORs and continuity-corrected ORs; the latter utilized a Haldane-Anscombe correction (adding a constant of 0.5 to all cells).^36^ P-values for both methods were derived using the Wald test based on the standard error of the log-odds.^37^

#### Statistical Analysis

Rare variants were analysed in functional categories. At gene-level, in three categories: (1) Non-synonymous (missense, in-frame indel, stop/start retained, protein altering, coding sequence, splice region); (2) CADD > 20 (top 1% predicted deleterious); and (3) Loss-of-Function (LoF) (stop gained, frameshift, start/stop lost, splice acceptor/donor, transcript ablation, exon loss). Domain-level analysis was limited to non-synonymous category variants.

Association testing utilized optimal sequence kernel association test (SKAT-O),^38^ adjusting for sex, age, and the first five principal components. Cohort results were meta-analysed via Stouffer’s Z-score method,^39^ weighted by case counts. The meta-analysis was performed separately for (1) a limited meta-analysis with AMP-PD and UKBB cases only and with limited controls and (2) a full meta-analysis with AMP-PD and UKBB with proxy. False Discovery Rate (FDR) corrections^40^ were applied separately for gene- and domain-level tests.

## Results

### Machine Learning Prioritizes 406 Genes for Parkinson’s Disease Association

To capture a comprehensive and high-yield map of the genetic architecture underlying PD, we constructed a composite set of risk loci by integrating the two largest available GWAS meta-analyses. We incorporated summary statistics from the recent European GP2 study and manually reinstated unique loci from the previous meta-analysis that were absent in the newer release, resulting in a final dataset of 147 unique PD risk loci. To ensure broad capture of potential causal candidates, all protein-coding genes located within a +/- 1mb window of the independent lead variants within each locus were extracted for prioritization.

We prioritized the top three scoring genes in each locus. This yielded a total of 406 unique genes prioritized across the 147 risk loci (See Supplementary Table 3 and Figure 2). 23 genes were prioritized multiple times due to overlapping neighbouring loci. Notable overlaps included *LSM7*, *SPPL2B*, and *TMPRSS9*, which were prioritized in three separate loci (122, 146, and 147). Other recurrent prioritizations included *BIRC6*, *SDHC*, *MED12L*, *AS3MT*, *TOX3*, *ADARB1*, *IQCB1*, and *ZSCAN31*, each emerging as the top-ranked prioritized gene in paired loci. We assessed the distribution of probability scores among the 406 prioritized genes. A high probability score (>0.75) was assigned to 33 genes (8.1%), including more established genes such as *SNCA* (0.86), *LRRK2* (0.86), *GCH1* (0.84), and *CTSB* (0.91) as well as prioritized genes like *ZYG11B* (0.92), *PXK* (0.92), *LMBRD1* (0.93). A further 142 genes (35.0%) received moderate probability scores between 0.50 and 0.75.

**Figure 2.**
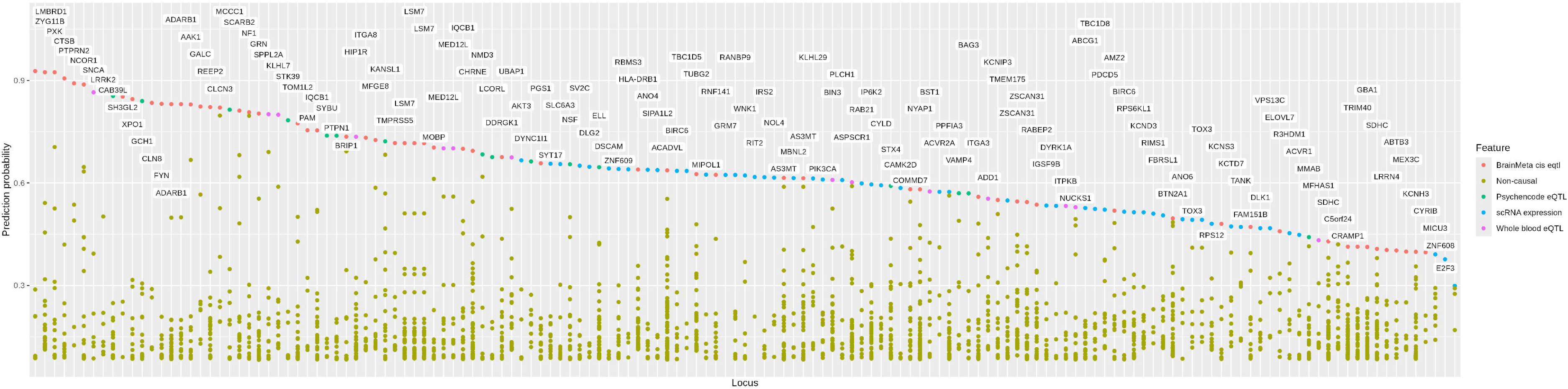
Probability scores of the Parkinson’s disease genome-wide association study candidate genes. This figure shows the probability scores from the machine learning model for each locus in the Parkinson’s disease genome-wide association study loci sorted in descending order. For each gene, the top non-distance feature was used to colour the data.

Conversely, the majority of the prioritized list showed lower confidence, with 231 genes (56.9%) scoring below 0.50. Notably, 123 genes (30.3%) had scores <0.25, indicating that for many loci, even the second or third-ranked prioritized gene had weak predictive evidence compared to the top nomination. For example, while *CLCN3* (0.82) and *MFAP3L* (0.80) in locus 43 both showed strong support, other loci such as 51 (*E2F3*, 0.29) and 131 (*DSCAM*, 0.64; *MX1*, 0.10) showed a steep drop-off in probability after the primary prioritized gene (See Supplementary Table 3).

### Feature Contribution and Predictive Drivers

To interpret the machine learning model, we utilized Shapley Additive exPlanations (SHAP)^41^ values to quantify the contribution of each feature to the final probability score. The instance-level contribution of these features is visualized in Figure 3, while the global hierarchy of feature importance is summarized in Figure 4.

**Figure 3.**
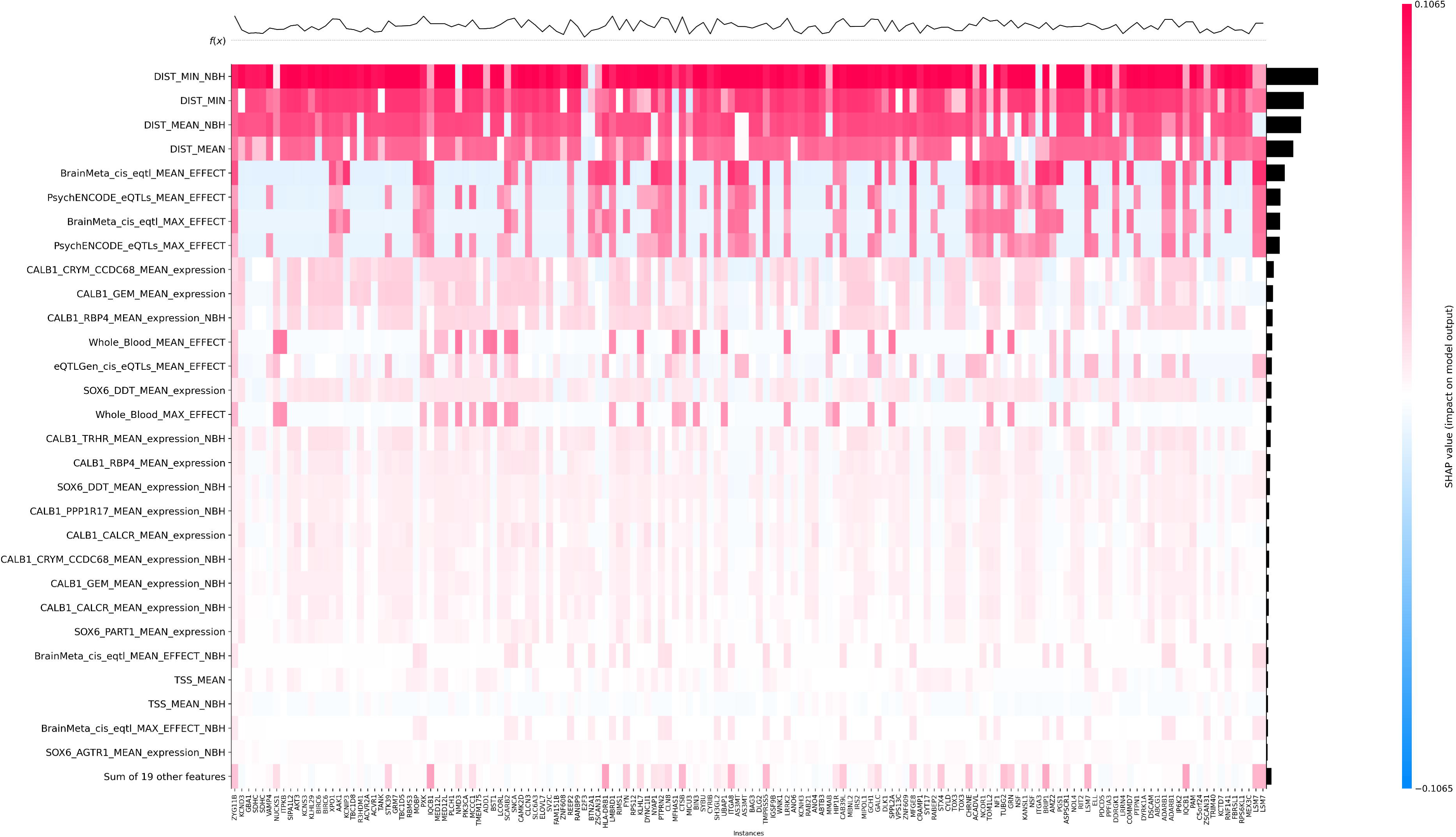
Heat map of feature importance. The heat map is generated using Shapley Additive exPlanations (SHAP) value for the top candidate gene in each locus. The plot at the top represents the probability score of each gene.

**Figure 4.**
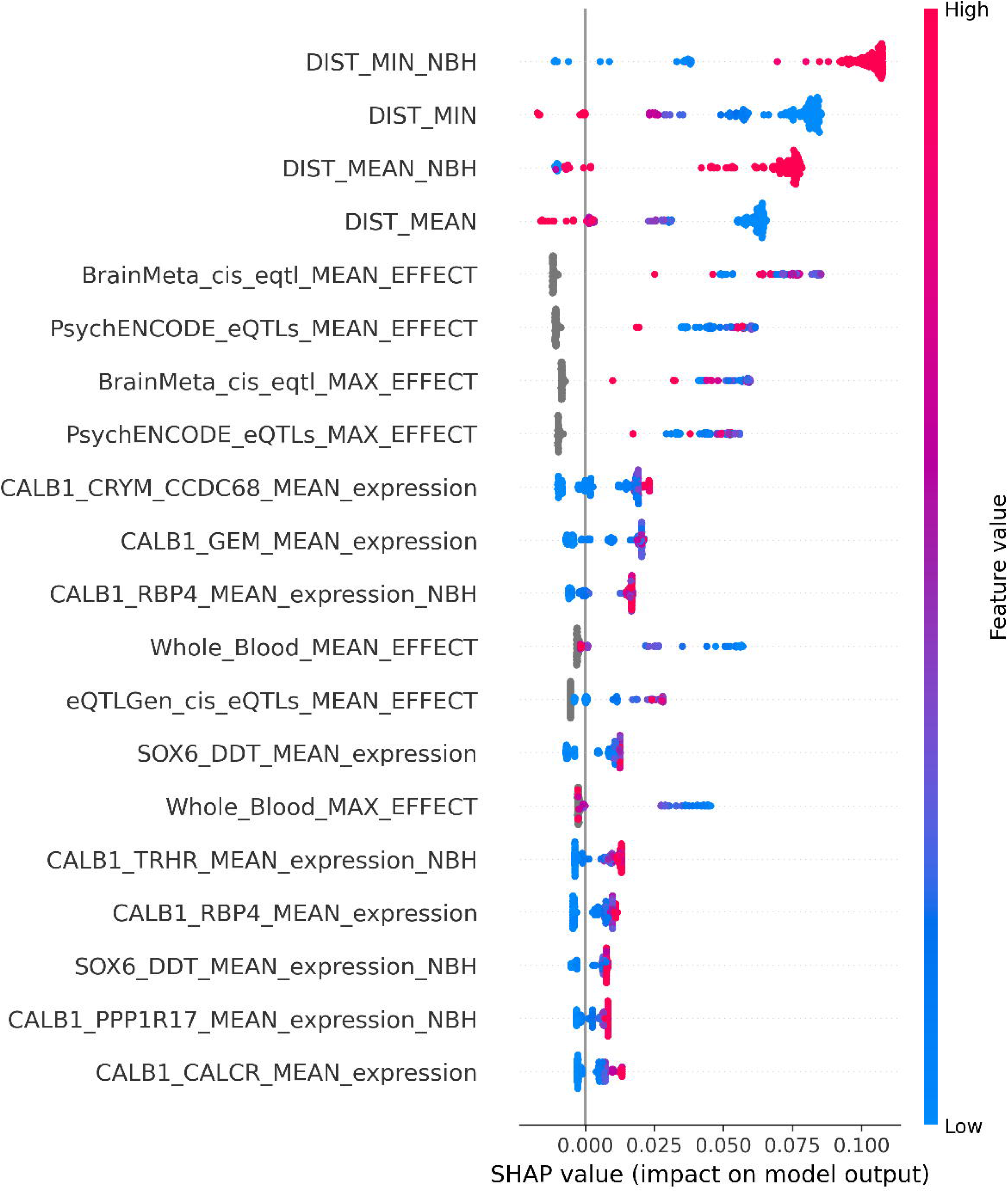
Feature importance for the Parkinson’s disease genome-wide association study gene prioritization model. Bee-swarm plot of feature importance using Shapley Additive exPlanations values along with the distribution of genes based on feature value.

Consistent with established GWAS biology,^12^ genomic distance features (e.g., DIST_MIN, DIST_MEAN) were the dominant drivers of the model. As illustrated in the top rows of Figure 3, proximity to the locus risk variants provided a high baseline probability for the majority of prioritized genes. However, the model demonstrated the capacity to override physical proximity when functional evidence was sufficiently strong. In 27 of the 147 risk loci (18.4%), the top-ranked candidate was not the gene with the lowest mean distance to the risk variants (See Supplementary Table 3). Beyond this distance-based prior, the model identified specific functional genomic signals as critical discriminators. As shown in Figure 4, the next most predictive features were cis-eQTL effect sizes derived from large-scale brain meta-analyses (BrainMeta) and the PsychENCODE consortium. This indicates that when genes are equidistant from a risk variant, the model prioritizes the gene with strong regulatory evidence in relevant tissue types. Additionally, we observed a distinct, high-specificity contribution from single-cell gene expression profiles. The model prioritized expression within specific *CALB1*-positive neuronal subpopulations (e.g., CALB1_CRYM_CCDC68 and CALB1_GEM). While these features had a lower global impact than distance, Figure 3 demonstrates that they contributed strongly to the scoring of specific gene subsets, suggesting they capture cell-type-specific vulnerabilities that broad tissue expression metrics miss.

### Burden analyses reveal novel targets at both gene- and domain-level

To further validate the prioritized genes and to identify novel rare variant burden associations, we performed rare variant burden analyses via SKAT-O in AMP-PD, UKBB, and UKBB with proxy cohorts (Table 1). The analysis included the 406 genes prioritized via our machine-learning prioritization framework and 767 domain-level targets identified by our domain demarcation process (See Supplementary Table 4). Post quality control steps, the analysis included a total of 27,777 unique rare variants from AMP-PD, 49,177 from UKBB, 155,399 from UKBB with proxy.

We subsequently performed two meta-analyses: (1) a limited meta-analysis with AMP-PD and UKBB (without proxies) and (2) a full meta-analysis with AMP-PD and UKBB (including proxies). The complete list of burden results is detailed in the Supplementary Tables 5 and 6 for gene- and domain-level analyses, respectively. The complete list of variants with detailed functional annotations, population frequencies, and OR calculations have been made accessible at: https://doi.org/10.5281/zenodo.18285356. The subset of findings reaching nominal significance (P < 0.01) in the full meta-analysis is presented in Tables 2 (gene-level) and 3 (domain-level). For these prioritized targets, detailed variant-level statistics across all functional categories are stratified by cohort in Supplementary Tables 7–9 (gene-level) and Supplementary Tables 10–12 (domain-level).

**Table 2.** Gene-level rare variant burden analysis results nominally significant (*P* < 0.01) in the full meta-analysis.

| **Gene** | **Variant Type** | **AMP-PD**<br>**$P_{FDR}$** | **UKBB**<br>**$P_{FDR}$** | **Limited Meta-analysis**<br>**$P_{FDR}$** | **UKBB with proxy**<br>**$P_{FDR}$** | **Full Meta-analysis**<br>**$P_{FDR}$** |
| --- | --- | --- | --- | --- | --- | --- |
| *GBA1* | CADD>20 | 0.07 | $6.55 \times 10^{-9}$ | $1.70 \times 10^{-10}$ | $3.68 \times 10^{-17}$ | $9.94 \times 10^{-20}$ |
| *GBA1* | Non-synonymous | 0.64 | $1.61 \times 10^{-18}$ | $4.34 \times 10^{-14}$ | $6.81 \times 10^{-15}$ | $1.25 \times 10^{-15}$ |
| *GBA1* | LoF | 0.66 | 0.97 | 0.82 | 0.02 | $7.65 \times 10^{-3}$ |
| *ANKRD27* | CADD>20 | 0.98 | 0.36 | 0.76 | $2.26 \times 10^{-3}$ | 0.03 |
| *UBXN2A* | CADD>20 | 0.95 | 0.59 | 0.81 | 0.02 | 0.04 |
| *ERCC8* | LoF | 0.84 | 0.59 | 0.68 | 0.06 | 0.04 |
| *FAM171A1* | LoF | 0.94 | 0.59 | 0.76 | 0.02 | 0.04 |
| *LRRK2* | CADD>20 | 0.92 | 0.59 | 0.81 | 0.03 | 0.06 |
| *LRRK2* | Non-synonymous | 0.97 | 0.59 | 0.88 | 0.02 | 0.12 |
| *UBXN2A* | Non-synonymous | 0.95 | 0.59 | 0.81 | 0.07 | 0.18 |
| *ADNP* | Non-synonymous | 0.95 | 0.85 | 0.88 | 0.11 | 0.24 |
| *LRRC45* | LoF | $6.28 \times 10^{-4}$ | 0.87 | 0.08 | 0.99 | 0.28 |
| *BNC2* | CADD>20 | 0.04 | 0.59 | $7.69 \times 10^{-3}$ | 0.99 | 0.28 |
| *CACUL1* | Non-synonymous | 0.70 | 0.95 | 0.81 | 0.85 | 0.48 |
| *KCNMB3* | CADD>20 | 0.81 | 0.88 | 0.76 | 0.83 | 0.52 |
| *SNCA* | Non-synonymous | $2.67 \times 10^{-3}$ | 0.95 | 0.37 | 0.99 | 0.52 |
| *MTMR7* | CADD>20 | 0.84 | 0.67 | 0.76 | 0.74 | 0.52 |
| *NCAM1* | CADD>20 | 0.90 | 0.80 | 0.81 | 0.57 | 0.52 |

AMP-PD – Accelerating Medicines Partnership – Parkinson Disease, UKBB – UK biobank, CADD – Combined Annotation Dependent Depletion.
Meta-analysis definitions: (1) Limited: AMP-PD + UKBB PD cases (ICD, self-report, death records; N = 6,435 cases, 37,504 controls); (2) Full: AMP-PD + UKBB PD cases and proxies (N = 20,324 cases, 343,160 controls).

**Table 3.** Domain-level rare variant burden analysis results nominally significant (*P* < 0.01) in the full meta-analysis.

| Gene | Domain | Variant Type | AMP-PD<br>$P_{FDR}$ | UKBB<br>$P_{FDR}$ | Limited Meta-analysis<br>$P_{FDR}$ | UKBB with proxy<br>$P_{FDR}$ | Full Meta-analysis<br>$P_{FDR}$ |
| --- | --- | --- | --- | --- | --- | --- | --- |
| *GBA1* | Glycosyl hydrolase family 30 TIM-barrel domain | Non-synonymous | 0.95 | $1.13 \times 10^{-15}$ | $3.40 \times 10^{-12}$ | $3.02 \times 10^{-12}$ | $5.14 \times 10^{-13}$ |
| *LRRK2* | Protein kinase domain | Non-synonymous | 0.95 | 0.59 | 0.83 | $9.68 \times 10^{-12}$ | $3.01 \times 10^{-10}$ |
| *GBA1* | Glycosyl hydrolase family 30 beta sandwich domain | Non-synonymous | 0.95 | $2.10 \times 10^{-4}$ | $1.93 \times 10^{-3}$ | $3.40 \times 10^{-5}$ | $1.88 \times 10^{-5}$ |
| *ADNP* | ADNP zinc finger domain | Non-synonymous | 0.95 | 0.86 | 0.87 | $3.23 \times 10^{-3}$ | $7.88 \times 10^{-3}$ |
| *NOL7* | U3 small nucleolar RNA-associated protein NOL7 C-terminal | Non-synonymous | 0.95 | 0.70 | 0.83 | 0.38 | 0.29 |
| *TOX3* | High mobility group box domain | Non-synonymous | 0.95 | 0.86 | 0.83 | 0.38 | 0.29 |
| *BNC2* | Zinc finger C2H2-type | Non-synonymous | 0.95 | 0.81 | 0.83 | 0.45 | 0.40 |
| *UBXN2A* | SEP domain | Non-synonymous | 0.95 | 0.59 | 0.83 | 0.37 | 0.46 |
| *KLHL29* | BTB/Kelch-associated | Non-synonymous | 0.95 | 0.59 | 0.83 | 0.45 | 0.71 |
| *SNRNP70* | RNA recognition motif domain | Non-synonymous | 0.95 | 0.94 | 0.83 | 0.99 | 0.71 |

AMP-PD – Accelerating Medicines Partnership – Parkinson Disease, UKBB – UK biobank.
Meta-analysis definitions: (1) Limited: AMP-PD + UKBB PD cases (ICD, self-report, death records; N = 6,435 cases, 37,504 controls); (2) Full: AMP-PD + UKBB PD cases and proxies (N = 20,324 cases, 343,160 controls).

At gene-level, post-corrections, we identified significant associations (*P_FDR_* < 0.05) in *GBA1* and *BNC2* in the limited meta-analysis, *GBA1*, *ANKRD27*, *FAM171A1*, *ERCC8*, and *UBXN2A* in the full meta-analysis, and *SNCA* and *LRCC45* genes in AMP-PD, which did not survive either meta-analysis. As expected, rare variants within *GBA1* and *SNCA*, two well-established PD risk genes showed significant associations: in the case of *GBA1,* the signal was across all three functional categories (non-synonymous, CADD > 20, and LoF) in the full meta-analysis, whereas, for the *SNCA* gene, the signal was limited to the AMP-PD cohort and the non-synonymous rare variants.

None of the novel genes identified in this analysis were the top-ranked candidate in their respective loci. *BNC2* (Locus 70) was prioritized as the third candidate. *ANKRD27* (Locus 124), *FAM171A1* (Locus 72), and *ERCC8* (Locus 45) were all ranked second in their respective loci, while *UBXN2A* (Locus 12) was the third candidate. Finally, *LRRC45* (Locus 119) was the second-highest ranked gene. Specific probability scores for each candidate are listed in Supplementary Table 3.

The sole novel signal in the limited meta-analysis, observed in *BNC2* (for variants with CADD score > 20), was driven by a significant cohort-level association in AMP-PD. This signal involved 530 variants, mostly intronic, with no single variant emerging as a clear driver (See Supplementary Table 7), therefore this association should be considered with caution.

Among the novel hits from the full-meta-analysis, *ANKRD27* (Full meta-analysis *P_FDR_* = 0.027 in the CADD > 20 category) was driven primarily by a single recurrent missense variant, p.Arg21Cys, which showed a consistent risk effect across all three cohorts (UKBB with proxy: OR = 1.20, 95% CI: 1.08–1.34, *P* = 1.03 x 10^-3^; UKBB: OR = 1.29, 95% CI: 1.01–1.65, *P* =0.04; AMP-PD: OR = 1.21, 95% CI: 0.81–1.80, *P* = 0.36). The remaining three gene-level hits in the full-meta-analysis (*FAM171A1* and *ERCC8* in the LoF category; *UBXN2A* in the CADD > 20 category) did not implicate specific driver variants, appearing instead to result from the cumulative burden of multiple rare alleles (See Supplementary Tables 7–9).

In the case of *LRRC45*, significant signals were limited to the AMP-PD cohort in both the CADD > 20 (*P_FDR_*= 6.28×10^-4^) and LoF (*P_FDR_* = 5.01×10^-3^) categories. Both signals were driven by the same potentially protective frameshift variant p.Met607ArgfsTer57 found in 3 cases and 31 controls in AMP-PD (OR = 0.11, 95% CI: 0.04–0.38, *P* = 3.60 x 10^-4^). This variant was not present in the UKBB cohort, which explains the limitation of signal only to AMP-PD.

At domain-level, post-corrections, non-synonymous rare variants in the *GBA1* 30 TIM-barrel and 30 beta-sandwich domains were found significantly associated, overlapping with the gene-level results. Furthermore, the *LRRK2* Kinase domain rare non-synonymous variants showed post-FDR significant association, which was not captured at gene-level for *LRRK2* (Variant level associations detailed in Supplementary Tables 10–12). Additionally, we identified a novel, significant domain-specific association in *ADNP* within its Zinc Finger domain. Importantly, *ADNP* did not reach significance in the gene-level burden test, demonstrating the ability of our domain-demarcation pipeline to dissect the structural localization of this burden. In the machine learning prioritization, *ADNP* was the second-ranked candidate in its locus, scoring below the top hit *PTPN1.* No clear variant-level association could be discerned as the association seemed to be driven by a cumulative effect of multiple variants within the domain (See Supplementary Tables 10–12).

## Discussion

In this study, we leveraged the latest and largest European GWAS meta-analysis to systematically prioritize and interrogate the genetic architecture of PD. Starting from 147 unique risk loci, our machine-learning framework prioritized 406 candidate genes. We identified several recurrent nominations across adjacent loci (e.g., *LSM7*, *BIRC6*, *TOX3*), implying that these regions likely represent single risk signals driven by extensive linkage disequilibrium rather than independent associations. While our machine learning model relied heavily on genomic distance, it importantly integrated functional genomic features, such as brain-specific eQTLs and cell-type-specific expression, identifying candidates that were not always the closest to risk variants in the locus.

Through our rare variant burden analysis, which included the demarcation of 406 candidate genes into 767 functional domains, we verified seven novel genes prioritized by machine learning, one of which was validated exclusively through domain-level resolution. This complementary approach proved critical for these findings as the seven genes were prioritized as second and third candidates respectively based on the machine learning model. By using the framework to define a broad search space rather than relying on a single high-confidence prediction, we enabled the rare variant burden analysis to identify risk associations that would have been overlooked by the prioritization model alone. In the gene-level analysis, we identified robust associations in *BNC2* in the limited-meta-analysis and in *ANKRD27, FAM171A1, ERCC8*, and *UBXN2A* within the full meta-analysis. The signal in *BNC2* was mostly driven by intronic variants with high CADD scores exclusively in the AMP-PD cohort. Given the lack of replication and the non-coding nature of the drivers, it remains unclear if this signal reflects *BNC2* biology or regulatory effects on neighboring genes, necessitating caution in interpretation.

The association in *ANKRD27* is particularly compelling given its known role in endosomal trafficking and interaction with the retromer complex, a pathway heavily implicated in PD pathology containing a PD-associated *VPS35* gene.^42^ This signal was driven by the recurrent missense variant p.Arg21Cys, which showed a consistent risk effect across all cohorts. Notably, a non-coding variant within *ANKRD27* (rs11672605) shows genome-wide significance (1.61 x10^-8^) in European GWAS^5^ but is in high linkage disequilibrium (r^2^ > 0.8) with the top locus hit (rs11669800), making it difficult to distinguish the causal driver via GWAS alone. Our rare variant finding suggests that *ANKRD27* and specifically the p.Arg21Cys variant may be the functional drivers of this locus.

The associations in *FAM171A1* and *ERCC8* were driven largely by LoF variants. *ERCC8* encodes a component of the ubiquitin ligase complex involved in transcription-coupled nucleotide excision repair, and its dysfunction causes Cockayne syndrome, a rare segmental progeroid disorder characterized by growth failure, severe photosensitivity, and progressive neurodegeneration.^43^ Given the accumulation of DNA damage and mitochondrial dysfunction in PD, *ERCC8* represents a plausible link to genomic instability. *FAM171A1* offers a plausible mechanistic link to PD pathology; recent evidence suggests that along with its paralog *FAM171A2*, *FAM171A1* may regulate the levels of progranulin (*GRN*), a recognized PD candidate gene.^44^ The role of *UBXN2A*, a ubiquitin-binding protein involved in proteasomal degradation,^45^ remains less established in the context of neurodegeneration. As the signals concerning these novel genes were cumulative rather than variant-specific, functional studies targeting the expression of these genes are warranted to elucidate their role in PD pathobiology.

Conversely, our analysis of *LRRC45* in the AMP-PD cohort revealed a potentially protective LoF signal driven by the frameshift variant p.Met607ArgfsTer57. *LRRC45* is a centrosomal protein involved in ciliogenesis.^46^ Since PD pathology has been linked to defects in primary cilia and centrosomal cohesion (notably involving *LRRK2*),^47^ a protective LoF effect could imply that reduced *LRRC45* activity mitigates specific cellular deficits, or that the gene normally exerts a deleterious gain-of-function in the disease state.

Our results emphasize that functional specificity via domain-level analysis can uncover insights masked by global gene-level tests. A prime example is *LRRK2*; consistent with literature showing that the p.G2019S variant exerts its effect via the kinase domain,^4,48^ our analysis successfully isolated the burden to this specific domain. Parallel to this, we identified a novel domain-specific association in *ADNP* restricted to its Zinc Finger domain. *ADNP* is a critical neuroprotective transcription factor.^49^ Crucially, *ADNP* expression is reduced in the substantia nigra of PD patients, where it functions alongside the aging-regulator *SIRT1* to orchestrate microtubule dynamics and histone methylation.^50^ The zinc finger domain burden identified here may therefore compromise these neuroprotective interactions, potentially mimicking the somatic *ADNP* mutations observed in Alzheimer’s disease that drive cytoskeletal impairment.^51^ While it failed to reach significance at the gene level, the domain-resolved signal highlights the power of this approach to filter out protein-wide noise and pinpoint potentially risk-influencing motifs.

Our study has several limitations. First, both the GWAS priors and rare variant burden analyses were restricted to populations of European ancestry. Larger, diverse datasets are needed for more global conclusions. Second, while quality control was rigorous, slight differences between the AMP-PD and UKBB pipelines may introduce minor heterogeneity. Third, our domain demarcation relied on consensus databases (Pfam, SMART, PROSITE) which utilize predictive algorithms to define functional boundaries. Although we sought to minimize inconsistencies by anchoring our pipeline to the validated InterPro standard, this predictive approach implies that certain domains may be missed or may not strictly align with manually curated definitions found in prior literature. Finally, the inclusion of proxy cases in the UKBB meta-analysis warrants restraint in interpretation, as proxy-based phenotypes share only half the genetic liability of clinically diagnosed PD.

In conclusion, our study shows that integrating machine learning-based gene prioritization with domain-resolution burden analysis offers a robust framework for dissecting the complex genetic architecture of PD. By prioritizing candidates based on multi-omic features and proximity to non-coding GWAS hits, we successfully identified novel risk genes *BNC2*, *ANKRD27*, *FAM171A1*, *ERCC8*, and *LRRC45* and uncovered a domain-level signal in *ADNP* (Zinc Finger) that was masked at the gene level. These findings not only highlight specific biological processes, such as endosomal trafficking, DNA repair, and ciliary function, but also demonstrate that multi-omics based, high-resolution genomic interrogation is highly valuable for translating GWAS loci into actionable biological targets. Future experimental validation of these candidates is critical to fully decipher their mechanistic roles in disease pathogenesis.

## Data availability

The WGS cohorts accessed and analysed in this study for the purposes of the burden analysis are accessible (conditional to institutional approval) at: AMP-PD Knowledge Platform (https://www.amp-pd.org) and the UK Biobank Research Analysis Platform. All information directly relevant to the study is included in the text and the supplementary files of this article, except for the full range of analysed variants, which have been made accessible online at: https://doi.org/10.5281/zenodo.18285356.

## Code availability

All the scripts used in the machine learning prioritization and burden analyses are shared at the GitHub repository link: https://github.com/gan-orlab/ML_and_Burden_PD.

## Acknowledgements

We extend our deepest gratitude to the participants from all cohorts included in this study for their essential contributions to this work. This research was performed using the NeuroHub infrastructure and was supported in part by the Canada First Research Excellence Fund, awarded to McGill University for the Healthy Brains, Healthy Lives (HBHL) initiative, as well as by Calcul Québec and the Digital Research Alliance of Canada. This study also made use of data from the UK Biobank under Application Number 45551.

Data for this article were obtained on 10 June 2025 from the Accelerating Medicines Partnership® (AMP®) Parkinson’s Disease (AMP PD) Knowledge Platform, release version 4.2. Further details on AMP-PD are available at https://www.amp-pd.org. The AMP® PD program operates as a public-private partnership managed by the Foundation for the National Institutes of Health (FNIH) and funded by the National Institute of Neurological Disorders and Stroke (NINDS) in collaboration with the Aligning Science Across Parkinson’s (ASAP) initiative. Industry partners include Celgene Corporation (a subsidiary of Bristol-Myers Squibb Company), GlaxoSmithKline plc (GSK), The Michael J. Fox Foundation for Parkinson’s Research (MJFF), Pfizer Inc., AbbVie Inc., Sanofi US Services Inc., and Verily Life Sciences. “ACCELERATING MEDICINES PARTNERSHIP” and “AMP” are registered service marks of the U.S. Department of Health and Human Services.

The clinical data and biosamples used in this article were derived from the following cohorts: (i) the BioFIND study (MJFF and NINDS); (ii) the Harvard Biomarkers Study (HBS) and the Stephen & Denise Adams Center for Parkinson’s Disease Research at Yale School of Medicine (CPDR-Y); (iii) the National Institute on Aging (NIA) International Lewy Body Dementia Genetics Consortium Genome Sequencing in Lewy Body Dementia Case-control Cohort (LBD); (iv) the MJFF LRRK2 Cohort Consortium (LCC); (v) the NINDS Parkinson’s Disease Biomarkers Program (PDBP); (vi) the MJFF Parkinson’s Progression Markers Initiative (PPMI); (vii) the NINDS Study of Isradipine as a Disease-modifying Agent in Subjects With Early Parkinson Disease, Phase 3 (STEADY-PD3); and (viii) the NINDS Study of Urate Elevation in Parkinson’s Disease, Phase 3 (SURE-PD3).

BioFIND is sponsored by MJFF with support from NINDS; current study information is available at https://michaeljfox.org/news/biofind. The BioFIND investigators did not participate in the review of this manuscript’s data analysis or content. Genome sequence data for the Lewy body dementia case-control cohort were generated at the U.S. National Institutes of Health Intramural Research Program, supported in part by the NIA (program #: 1ZIAAG000935) and NINDS (program #: 1ZIANS003154). The Harvard Biomarker Study (HBS) is a collaboration of HBS investigators (listed at https://www.bwhparkinsoncenter.org/biobank/) and is funded through philanthropy, NIH, and non-NIH sources. The Stephen & Denise Adams Center for Parkinson’s Disease Research of Yale School of Medicine is similarly funded through philanthropy, NIH, and non-NIH sources. Neither the HBS nor CPDR-Y investigators participated in reviewing the data analysis or content of this manuscript.

Data were obtained from the MJFF-sponsored LRRK2 Cohort Consortium (LCC). Current study information can be found at https://www.michaeljfox.org/biospecimens. The LCC investigators did not review the data analysis or manuscript content. PPMI is sponsored by MJFF and supported by a consortium of scientific partners (full list available at https://www.ppmi-info.org/about-ppmi/who-we-are/study-sponsors). For up-to-date study information, visit https://www.ppmi-info.org. PPMI investigators did not participate in the review of the data analysis or content of this manuscript. The Parkinson’s Disease Biomarker Program (PDBP) consortium is supported by NINDS at the NIH. A full list of PDBP investigators is available at https://pdbp.ninds.nih.gov/policy. PDBP investigators did not review the data analysis or content of this manuscript. STEADY-PD3 is funded by NINDS at the NIH with support from MJFF and the Parkinson Study Group. Additional information is available at https://clinicaltrials.gov/ct2/show/study/NCT02168842. The STEADY-PD3 investigators did not review the data analysis or content of this manuscript. SURE-PD3 is funded by NINDS at the NIH with support from MJFF and the Parkinson Study Group. Additional information is available at https://clinicaltrials.gov/ct2/show/NCT02642393. The SURE-PD3 investigators did not review the data analysis or content of this manuscript.

## Author Contributions

Contributors S.C.P., E.Y., and Z.G.-O. conceived and designed the study. E.Y. developed the machine learning framework and performed the prioritization analysis. S.C.P. performed the domain demarcation and rare variant burden analysis and wrote the first draft of the manuscript. S.K. and M.Z. assisted with data extraction and quality control. All authors contributed to the interpretation of the data and critical revision of the manuscript. Z.G.-O. obtained funding and supervised the study. All authors had full access to all the data in the study and had final responsibility for the decision to submit for publication.

## Supplementary material

Supplementary material is provided in the supplementary tables.

## Abbreviations

AMP-PD: Accelerating Medicines Partnership for Parkinson’s Disease
eQTL: Expression quantitative trait loci
GWAS: Genome-wide association studies
PD: Parkinson’s disease
SKAT-O: Optimal sequenced kernel association test
UKBB: UK Biobank

